# Differences in pathogenetic mechanism between Tibetan and Han high-altitude polycythemia based on a whole genome-wide association study

**DOI:** 10.1101/2023.08.05.23293524

**Authors:** Basang Zhuoma, Zhang Shixuan, Ke Xianwei, Duoji Zhuoma, Yang La, Qiangba Danzeng, De Yang, Hu Zixin, Ma Yanyun, Hao Meng, Lin Zeshan, Li Yi, Wang Jiucun, Wu Juan

## Abstract

High-altitude polycythemia (HAPC) is a common and serious chronic disease affecting people of highland and plains ancestry living at high altitudes. This study investigated genetic susceptibility differences for HAPC among ethnic groups, with 2,248 volunteers participating, including 898 HAPC patients (n_Han_ = 450, n_Tibetan_ = 448). The study included a GWAS of 198 cases (n_Han_ = 100, n_Tibetan_ = 98), which revealed eight Tibetan HAPC-susceptibility single-nucleotide polymorphisms and four Han HAPC-susceptibility VARIANTs. Among them, the common polymorphism locus rs7618658 (*SNX4*, *P*_combine_<5e-8) was found and verified in both Tibetans and Han Chinese. Furthermore, the exploration of Tibetan *EPAS1* suggested that the rs1374749 locus of *EPAS1, along* with other locus-linked status changes, may be a prevalence factor for HAPC, and the GGTAC haplotype in which it is located is a risk haplotype for HAPC (P = 5.14E-9, OR = 1.85).The study showed significant genetic susceptibility to HAPC between Tibetans and Han Chinese, with Tibetans mainly susceptible in oxygen-sensing pathways such as *EPAS1*, which are associated with several phenotypes (Hemoglobin, Hematocrit, and Platelets, etc), while Han Chinese patients exhibited susceptibilities mainly in cell differentiation and angiogenesis, such as in *SNX4* and *LPAR1*, which are similarly significantly associated with hemoglobin, hematocrit, and platelets.

## Introduction

High-altitude polycythemia (HAPC) is characterized by polycythemia^1^ (women: Hb ≥ 19 g/d L; men: Hb ≥ 21 g/d L) and significant increases in lung volume, thoracic cavity, hemoglobin levels, and hematocrit^1^ leading to increased blood viscosity and potential harmful consequences such as microcirculation dysfunction, vascular thrombosis, sleep disturbance and even death^1, 2^. HAPC is a unique altitude sickness with various complications including pulmonary vascular dysfunction^3^ and hypertension^4^, which may seriously affect the health of long-term and short-term residents at high altitudes. Epidemiological surveys have revealed significant difference in HAPC incidence rate among different ethnic groups. For example, the incidence rate is approximately 2.39% among Tibetan populations^5, 6^, 15% among people living in the Andes and 17.8% among Han Chinese people^7^. The increased susceptibility to HAPC from Tibetans to Andean populations to Han Chinese people suggests an intrinsic association between high-altitude acclimatization and susceptibility to HAPC. Studies indicate that Tibetans and Han Chinese have significant genetic differences and that Tibetan populations adapted to high-altitude environments earlier in human history^8^. Therefore, dissimilarities in the mechanisms of HAPC production between the two groups, such as the degree of variation in endothelial PAS domain protein 1 (*EPAS1)*^9,10^.

Migrating Han Chinese or people of Tibetan descent who live in Tibet adapt to low-oxgen environment by increasing hemoglobin levels^11, 12^ and red blood cells^13^ to alleviate the lack of oxgen supply to tissues. The present study suggests that these changes may be regulated by the activation of the oxygen-sensing pathway (*HIF-1A*/*EPAS1*)^14, 15^ which leads to an increased inflammatory response^16^, decreased lipid metabolism^17, 18^ and increased angiogenesis^19^. Although increasing hemoglobin and red blood cells can alleviate the oxygen supply problem, some Han Chinese mobile populations may not be able to adapt to hypoxic environments due to inter-individual differences in oxygen demand. Numerous studies have investigated possible genetic susceptibility similarities and differences for HAPC of Han Chinese and Tibetan populations. For example, a previous study found that the genetic susceptibility of Han Chinese to HAPC was associated with variants in *EPAS1*^9, 20^, *PIK3CD*^21^, *COL4A3*^21^, and *CARD14*^22^, while Tibetans had different genetic susceptibilities, such as *EPAS1*^6, 23^, *PPP1R2P1*^33^, *PIK3CD*^24^, *GLUT1*^25^, and *ITGA6*^23^. However, exploring HAPC prevalence patterns based on smaller samples may have limitations^23^. Therefore, studying genome-wide differences in HAPC between Tibetan and Han populations based on larger samples will help to understand genetic susceptibility differences between populations and may lead to more precise treatment.

In this study, we conducted genome-wide scanning microarray typing on 98 Tibetan and 100 Han Chinese HAPC patients with healthy controls GWAS comparison, including 74 Tibetans and 1,076 Han Chinese. Futhermore, the study recruited 700 HAPC patients for independent validation of VARIANTs with significant differences in GWAS (n_Tibetan_=350; n_Han Chinese_=350), with 900 healthy controls. We then performed GWAS studies, HAPC blood phenotype differential analysis, independent replicate validation and linkage disequilibrium (LD) analysis. The present study identified and validated one common protective factor, rs7618658 (*SNX4*). Additionally, LD analyses revealed risk features of AACGT-linked haplotypes in Tibetan HAPC and these 5 loci were significantly correlated with blood parameters, such as hemoglobin, including rs1374749, rs10178633, rs11678465, rs1868092, rs11689694.

## Materials and methods

### Study population

The blood samples were taken from the Physical Examination Center, Fukang Hospital, Lhasa, Tibet. All participants signed informed consents. Passed strict inclusion and discharge standards **(Table 3)**.

#### Case

A total of 898 samples of male HAPC (Hemoglobin ≥ 21 g/dL ^24^) Tibetan and Han population were included in the study. Among them, 198 samples (Tibetan: n=98, Han: n=100) were genotyped by ‘Illumina Human Omni ZhongHua-8 Bead chip’ (Genome-wide Scan). In addition, 700 samples (Tibetan: n=350, Han: n=350) were verified for differential VARIANTs by VARIANTscan method. Refer **Table 4** for details of the Sample.

#### Control

A total of 2129 samples (Tibetan: n=566, Han: n=1563) of male normal Tibetan and Han population were included in the study. Among them, 1028 Han samples^25^ were genotyped by ‘Illumina Human Omni ZhongHua-8 Bead chip’ (Genome-wide Scan), 48 samples of Han nationality were taken from the 1000 Genomes Project (https://www.internationalgenome.org, Han Chinese in Beijing, HCB). In Tibetan samples, 38 by Next generation sequencing (NGS) and 36 by ‘Illumina chip’. In addition, 900 samples (Tibetan: n=450, Han: n=450) were verified for differential VARIANTs by VARIANTscan method. Refer **Table 4** for details of the Sample.

### VARIANT Discovery Stage

#### Genome-wide association study (GWAS)

Based on the original data of genotyping. First, we performed the positive and negative chain correction of the chip data and merged two group data (case & control). After that, data quality control (QC) was performed by Plink (v 1.90) ^26^ **(Fig S1 D)**. The QC contains the following content: deleted VARIANTs were missing in 5% of samples, call rate>95%, VARIANTs Minor allele frequency(MAF) is greater than 0.001, Hardy-Weinberg Equilibrium (HWE) was greater than 0.0001. Logistic regression method to analyze the difference of VARIANTs between the two groups, and P value witnessed the genetic structure through PCA (**Fig S1 D**). P value after the correction is basically in line with the normal distribution **(Fig S1 D)**. With the ‘Ensembl’ annotated database, the resulting VARIANTs were annotated by ANNOVAR ^24^ according to the locations.

#### Gene ontology analyze

The metascape online tool (http://metascape.org) was used to pathway enrichment analysis, which genes set of the difference VARIANTs (P adj <0.05, 5e-5, 8e-8) between Tibetans and Hans. The database sources used include: GO(BP), KEGG, Reactome Gene Sets, Wiki Pathways (p-value < 0.01, similarity score > 0.3, enrichment factor > 1.5). In addition, We obtain the hypoxia-related gene set (n=200, **Table S6**) from the hallmark gene set(H) of the MSigDB^27^ database, and performed Venn analysis with the difference VARIANTs of the two groups. Perform pathway enrichment analysis on the gene list of Venn analysis results.

#### VARIANT verification Stage (VARIANTscan)

In this study, using the Illumina VARIANTscan method, we identified significantly different VARIANT site of large sample. Plink tools was used for VARIANTs quality control (QC), and a Chi-Squared Test between case and control. We set P value < 0.05 to screen the differentially VARIANTs. In addition, we use the fisher function in the R(v 4.1) Package ’metap’ (https://github.com/cran/metap) to combine the P values of GWAS and VARIANTscan after considering the sample weights to generate a combined P value (Tibetan: case=448, control=566; Han: case=450, control=1563). and set P value < 5e-8 to screen the differentially VARIANTs. To explore the effect of variant types of successfully verified VARIANTs on gene expression, we used the normal population expression matrix of the GTEx database^28^ to eQTL analysis.

#### Gene-based and PPI Network

The study used GWAS association results for HAPC in Tibetans and Han Chinese, and gene-based association analysis was performed using plink software for VARIANT loci with p-values less than 0.01 to explore the overall effect of whole genes in differentiating HAPC patients. Next, PPI networks were constructed using gene-based significant genes (P<0.01) which included 347 genes for Tibetans and 11,271 genes for Han Chinese (Score>0.4). Subsequently, network centring using Cytoscape’s MCODE tool was performed to identify association modules between genes, and pathway enrichment studies of module genes were performed using GO and KEGG databases to explore the source of genetic susceptibility to HAPC in the two ethnic groups.

### Blood phenotype difference analysis

We collected 20 hematological phenotypes, including Hemoglobin (HGB), Mean hemoglobin concentration (MCHC), Erythrocyte (RBC), Hematocrit (PCV), Mean erythrocyte volume (MCV), Standard deviation (RDW-SD), Platelet compaction (PLT), Mean platele volume (MPV), Soterocyte (P-LCR), Lymphocyte (LYM), Lymphocyte% (LYM%), Monocyte (Mono), Monocyte% (Mono%), Neutrophile (NEUT), Neutrophile% (NEUT%), Basophilic granulocyte (BASO), Basophilic granulocyte% (BASO%), Eosnophils (EOS), Eosnophils% (EOS), Hematocrit (HTC), Neutrophile/lymphocyte(NLR), Hemameba (WBC).

A two-sided Wilcoxon signed-rank test was performed to compare the above phenotypes of HAPC and non-HAPC inside the Tibetan and Chinese Han, and the cutoff of P-value adopts multiple tests (0.05/20 = 0.0025). Then, Spearman correlation analysis was performed between VARIANTs in the 20kb region of EPAS1 within the Tibetan and Chinese Han population (a total of 17 sites in Tibetan and 9 sites in Han), to identify the VARIANT sites correlated with the above phenotypes (P value < 0.05, |Coeff| > 0.2). In addition, the construct of related networks in the phenotype with Spearman association analysis. (P value < 0.05, |Coeff| > 0.2).

### Relevant predictions of gene variants and protein structure functions

The study explored the protein structure and function of two Tibetan missense variant loci rs2307279 (A38P), rs2567705 (I611F) using multiple databases, AlphaFold^29^, I-Mutant2.0^30^, MutPred2^31^, PolyPhen-2^24^ for protein 3D folding structure, prediction of variant on protein stability, effect of known diseases, and deleterious variant prediction.

### Prediction model construction

#### Blood phenotypes for independent diagnosis of HAPC

The study used logistic regression to classify key phenotypes (13 phenotypes) in blood characteristics for HAPC separately and evaluated the model using a 5-fold cross-validation with AUC as the evaluation index^32–34^. A total of 791 cases were included in the Tibetan population and 751 cases were included in the Han population.

#### Nomogram analysis

The study used Alignment Diagram to quantify the predictors based on the integration of multiple predictors that were significantly associated with HAPC between Tibetan and Han Chinese based on a multifactorial regression analysis (logistic regression) using Nomogram. Consistency index (C-Index) and area under the characteristic curve (AUC), calculated by bootstrap, were used to assess the discrimination ability. c-Index and AUC values ranged from 0.5 to 1.0, where 0.5 indicated random chance and 1.0 indicated perfect fit. Typically, C-Index and AUC values greater than 0.7 indicate a reasonable estimate.

## Results

### HAPC-susceptible VARIANT sites

Genome-scale genotyping was conducted using blood samples of both HAPC patients and non-HAPC patients of Tibetan and Chinese Han ancestry. Approximately 771,178 VARIANTs in Tibetan samples and 540,348 VARIANTs in Chinese Han samples were preserved after population structure correction and VARIANT quality control. Compared with the control group, the Tibetan HAPCs had 77 VARIANTs (*P* < 5e-8; **Fig. 1A, Table S1**), including rs17801569 (*LINC01507*), rs7618658 (*SNX4*), rs2567705 (*COL27A1*), and rs2307279 (*PLA2G4C*). The Chinese Han HAPC patients had 25 (OR<1) VARIANTs (*P* <5e-8; **Fig. 1B, Table S1**), including rs17876027 (*F12*), rs1030005 (*LPAR1*), rs17801569 (*LINC01507*) and rs7618658 (*SNX4*). Significantly, rs17801569 (*LINC01507*) and rs7618658 (*SNX4*) are HAPC susceptibility VARIANTs common to both Tibetans and Han Chinese (**Fig. 1C**, *P* < 5e-8). The region with the highest distribution of the VARIANTs was intronic (**Fig. 1D**), with 26 VARIANTs in Tibetan people and 12 VARIANTs in Chinese Han people. There were 24 VARIANTs in the exon region of Tibetan people (**Fig. 1D**).

**Fig. 1.**
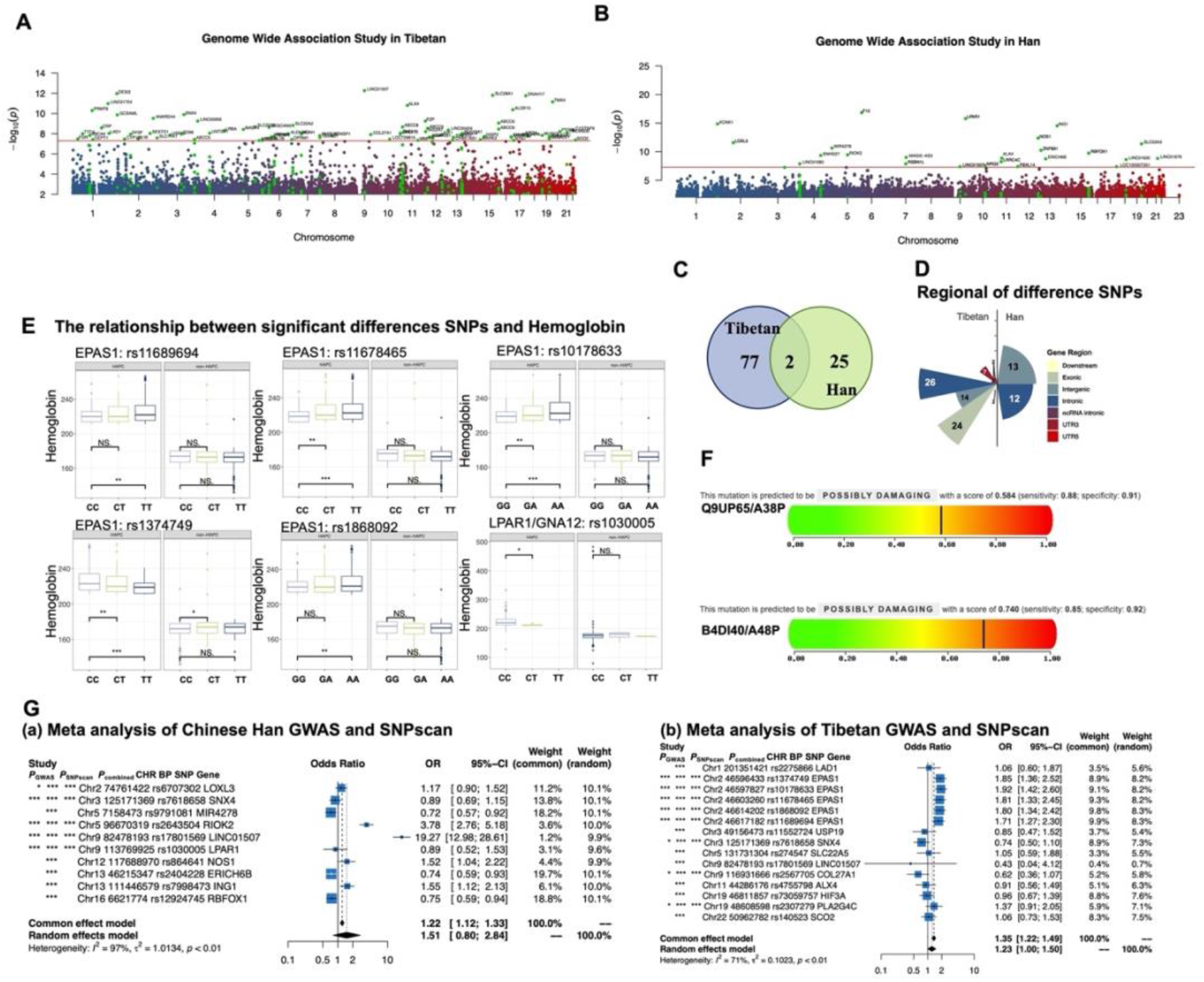
GWAS and Gene Base Study Results: **A.** GWAS analysis results in Tibetan. B: GWAS analysis results in Han Chinese. **B.** Analysis of Gene Base in Tibetan and Han Chinese. **C.** The shared VARIANTs of Tibetan and Han Chinese HAPC. **D.** Location area distribution of Tibetan and Han Chinese VARIANT. **E.** Association of significantly different VARIANTs with HGB in Tibetan and Han Chinese. **F.** PolyPhen-2 results suggest that rs2307279(missense) of Tibetan PLA2G4C increases protein structural stability. **(a).** PolyPhen-2 score. **(b).** The gamma (EC 3.1.1.4) region of pla2g4c protein is highly similar to protein B4DI40 in rs2307279 (A48P) with a prediction score of 0.740 for the protein. **(c)** Multiple sequence alignment of pla2g4c protein. **(d).** rs2307279(A48P) is located in the Alph fold structural threshold of the pla2g4c protein. **G.** Meta-analysis of the GWAS study and the VARIANTscan study.

Basis on GWAS analysis, 15 Tibetan VARIANTs and 10 Han VARIANTs **(Table S4)** were selected for independent and repeated validation according to the literature research on hypoxia and blood function or diseases and GWAS P-value. In a sample of 1,600 Tibetans and Han Chinese, the shared protective factor rs7618658 (*SNX4*, *P*_combine-Han_=3.21E-08; *P*_combine-Tibetan_=6.50E-09; **Table 1**) was successfully validated. Moreover, 5 *EPAS1* loci and 2 missense variants were verified in the Tibetan samples (*P*_combine_<5e-6, **Table 1**). This study also identified some VARIANTs that were significantly associated with HGB (*P*<0.01, **Fig. 1E**), including the TT genotype of rs1374749 (*EPAS1*), which reflected the lower occurrence of HGB in Tibetans; the GG genotype of rs2307279 (*PLA2G4C*), which reflected increased HGB; the CT genotype of rs1030005 (*LPAR1*/*GNA12*), which reflected the lower occurrence of HGB in Han Chinese. In addition, this work verified three HAPC protection factors in Han Chinese (OR<1, **Table 1**). Finally, meta-analysis was used to combine the validation population with the GWAS population, in which some of the mutant loci were significantly altered in OR due to the influence of the sample population (**Fig. 1G**).

To explore the candidate genes of HAPC, a Gene Based study was conducted based on the differential VARIANTs revealed by the GWAS (*P*_adj._ <0.01), which showed 348 genes in Tibetan samples and 734 genes in Han samples. Among these genes, 33 VARIANTs of *EPAS1* (**Table S5**) and 7 VARIANTs of *PLA2G4C* (**Table S5**) in Tibetan repeat-validated genes exhibited a significant difference in Gene Base. The *EPAS1* genetic variant was a feature of Tibetan evolution and participated in the regulation of red blood cell generation^35^. The rs1868092 (G>A, 3’-flanking) VARIANT was reported as a risk factor in a Han HAPC^36^. 4 VARIANTs detected in this study have not been reported to be related to HAPC, including rs1374749 (A>G, intron), rs10178633 (A>G, intron), rs11678465 (C>T, intron), and rs11689694 (T>C, 3’-flanking). In addition, in Han samples, 10 VARIANTs of the *LPAR1* gene (*P*= 1.56E-04, **Table S5**) and 1 VARIANT of the *GNA12* gene (*P*= 4.94E-03, **Table S5**) showed significant differences in Gene Base.

### Effect of mistranslated variant sites on protein structure

To determine whether variants found and validated in Tibetan versus Han Chinese populations have deleterious effects on protein structure and function, this study explored the risk scores of eight Tibetan VARIANTs and four Han Chinese VARIANTs using the PolyPhen-2 method. The results suggest the effect of two missense sites of Tibetan on protein structure (**Table 1**), including rs2307279 (A38P, *PLA2G4C*) and rs2567705 (*COL27A1*). The PolyPhen-2 results showed that the variant at the rs2307279 (A38P) locus led to the protein sequence being evaluated as POSSIBLY DAMAGING (Score1=0.584, Score2=0.740, **Table S7, Fig. 1F(a)(b)**), which suggests a possible effect of the variant on the protein function (**Fig. 1E**). Second, in the AlphaFold^29^ results the substitution of A and P position 38 was found to be essential for Alph folding (pLDDT=91.84). The I-Mutant2.0 results similarly suggested a significant increase in protein stability due to the rs2307279 (A38P) variant (RI=2). In the UK biobank database^27^, significant correlations (*P*= 2.1e-3, OR = 0.81) were found between the rs2307279 variant and disorders of lipid metabolism and coagulation.

### HAPC blood phenotype characteristic diagram

To explore the phenotypic differences between HAPC and non-HAPC in Han Chinese and Tibetan people, 20 blood phenotypes were collected. The Wilcox test (*P*<0.0021, **Fig. S1, Table 2**) showed that 13 phenotypes were significantly different between Tibetan-Han HAPC and non-HAPC samples. Blood phenotypic differences showed that the Tibetan and Han Chinese migrant HAPC populations had similar blood characteristics, which were mainly reflected in increased RBC, increased HCT, decreased PLT, increased RDW-SD, decreased MCHC and decreased LYM% (**Table 2**). Among them, RBC, MCHC, and LYM% were significantly different among HAPC (*P*< 0.05, **Fig. S1A**), while RBC was higher among Tibetans (**Table 2**), and LYM% and MCHC were higher among migrant Han Chinese (**Table 2**). In addition, RBC, HGB, and HTC were also significantly different among non-HAPC, and they were higher in migrant Han Chinese (*P*< 0.05, **Fig. S1A**). Considering the effect of the NEUT to LYM ratio (NLR) in hematocrit, it was found that the NLR would be significantly higher in the Tibetan HAPC population than in the Han population (**Fig. S1A, Table 2**). Different from the migrant Han Chinese, the Tibetan HAPC population showed an increase in WBC and a decrease in NEUT% (**Fig. 2A, Table 2**). In addition, MPV was significantly higher only in the Han migrant HAPC population (**Fig. 2A, Table 2**).

**Fig. 2.**
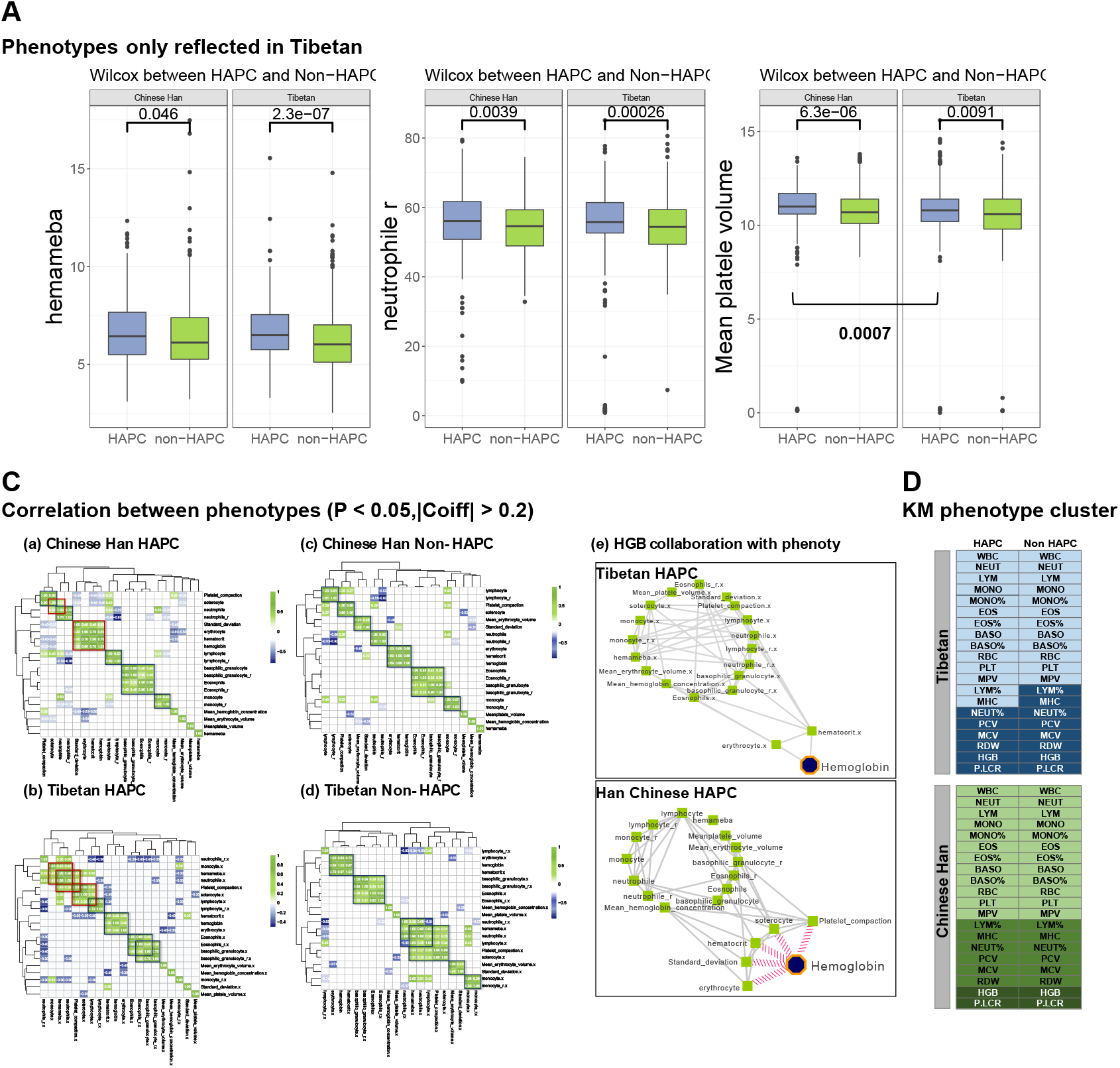
Blood phenotype difference analysis: **A.** Blood phenotype of significant differences only in Tibetan or Chinese Han HAPC and Non-HAPC. ***P< 0.0021 (0.05/23). **B**. Exploration of correlation between phenotypes and differentially validated VARIANTs between Tibetans and Han Chinese (P< 0.05). **C. (a)(b)(c)(d)** Spearman correlations between the four subgroups of Tibetans and Han Chinese. The red box in the figure represents the difference between HAPC and Non-HAPC. **(e).** Synergistic variation between HGB in HAPC population and phenotype between Tibetans and Han Chinese (using Spearman’s method). **D:** Blood phenotype KM cluster distribution.

Spearman’s network showed the difference in co-variation between Tibetan and Han blood phenotypes. Compared with the Tibetan non-HAPC group, the HAPC groups showed significant co-variation characteristics between WBC and PLT, P-LCR, LYM, and NEUT (*P*<0.0025, |Coeff|>0.2, **Fig. 2C(b)**). Compared with Han non-HAPC group, the HAPC group showed significant synergy between P-LCR and NEUT (*P*<0.0025, |Coeff|>0.2, **Fig. 2C(a)**), as well as increased synergy between RDW.SD and HGB, RBC, and HCT (*P*<0.0025, |Coeff|>0.2, **Fig. 2C(a)**). In addition, there were large phenotypic synergistic differences between Tibetan and Han HAPC populations, which were mainly reflected in the synergistic relationship between PLT and Mono and WBC in the Tibetan HAPC population (**Fig. 2C(a)(b)**), but not in Han Chinese. Finally, this work focused on the synergistic variation of HGB and phenotype differences between Tibetan and Han Chinese HAPC groups. Compared to the Tibetan HAPC population, the Han Chinese HAPC population had increased correlations of RDW.SD, PLT, and P-LCR with HGB (**Fig. 2C(e)**, *P*<0.05, |Coeff|>0.2). In addition, clustering studies using the KM algorithm revealed different patterns in the Tibetan HAPC population, which were shown by a change in the clustering pattern of LYM% with the other phenotypes (**Fig. 2D**).

Based on the blood phenotypic features of HAPC, this work focused on the possible correlations between the phenotypes and the discovered and validated VARIANTs. First, 9 blood phenotypes were shown to be correlated with the VARIANT locus of *EPAS1* in the Tibetan population (**Table S3**, P<0.05), including 2 differential phenotypes, NEUT% and WBC, which were unique to the Tibetan HAPC population. Second, *SNX4* (rs7618658) showed significant correlations with HGB, Mono, and LYM phenotypes in the Tibetan HAPC population (P<0.05). Among Chinese Han HAPC patients, this study found significant correlations between rs1030005 (*LPAR1*; *GNA12*) and 7 key phenotypes (**Table S3**, P<0.05), in which HGB, WBC, BOSO, EOS, RBC, and MCHC were significantly different between HAPC and non-HAPC patients (**Table S3**, P<0.0025).

### HAPC gene ontology exploration

To further identify the gene function of differential VARIANTs in Tibetan HAPC and Han HAPC patients, Gene Ontology (GO) enrichment analysis was conducted. Based on 77 VARIANTs in Tibetan patients (P_GWAS_<5e-8), 32 pathways were determined to be enriched (Counts_gene_>5, P<0.05, **Table S2, Fig. 3A(a)**), of which the top four most significantly enriched pathways were active transmembrane transporter activity, ABC transporters, arachidonic acid metabolic process and unsaturated fatty acid metabolic process. Based on 25 VARIANTs in Han Chinese patients, 75 pathways were determined to be enriched (Counts_gene_>2, P<0.05, **Table S2, Fig. 3A(b)**), of which the top most significantly enriched pathways were the adenylate cyclase-modulating G protein-coupled receptor signaling pathway and regulation of cell morphogenesis, including *GNA12*, *LPAR1*, *NOS1*, and other genes.

**Fig. 3.**
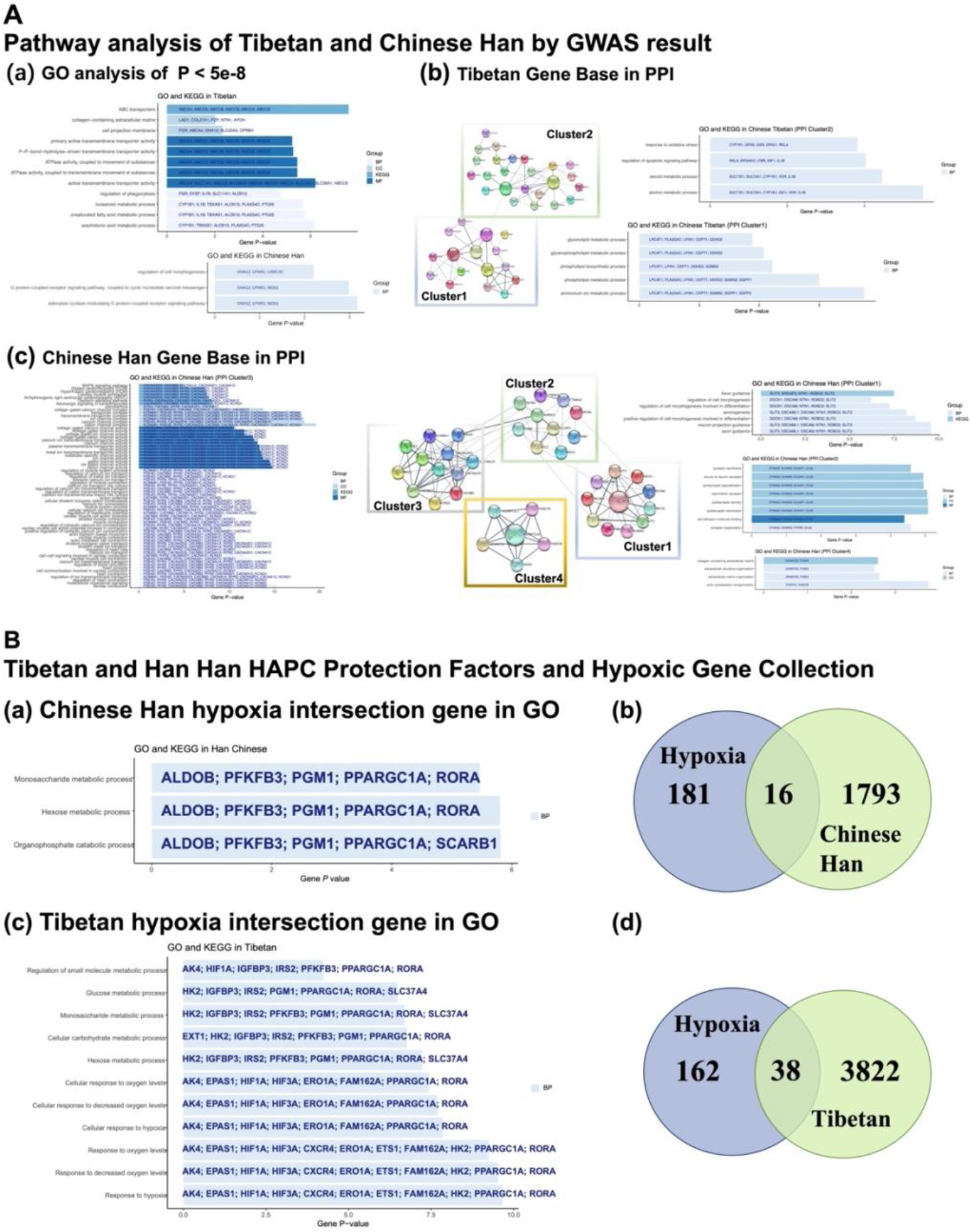
Genetic Ontology Exploration of Tibetan and Han HAPC Protection Factors: **A.** GO, KEGG analysis of HAPC in Chinese Han and Tibetan. **B. (a) Chinese Han** GO enrichment analysis. **(c)** Tibetan GO enrichment analysis. **(b)(d)** Analysis of Difference Gene VENN Analysis of Hypoxia Gene Collection and Tibetan and Han people.

**Fig. 4.**
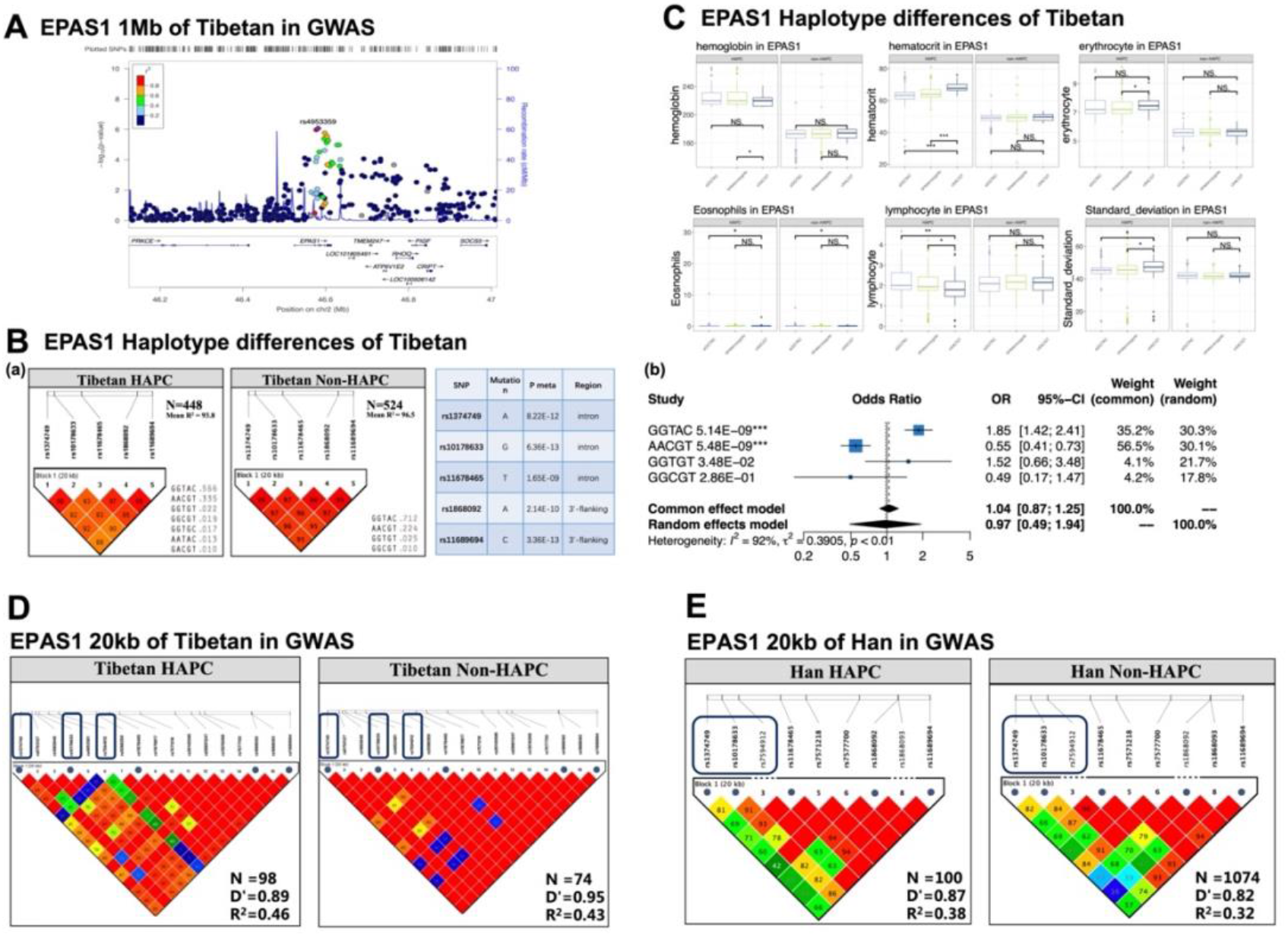
Exploration of EPAS1 and HAPC. **A:** Locus Zoom Analysis shows 500 MB region upstream and downstream of the EPAS1 gene. **B: (a)** LD linkage disequilibrium analysis of five EPAS1 VARIANTs in HAPC and Non-HAPC **(b).** Five VARIANT composition of multiple forest maps. **C.** The difference between the phenotype in the EPAS1 polarization type. **D.** LD linkage study within 20 kb of the region where the five EPAS1 genes are located in Tibetan GWAS. **E.** LD linkage study within 20 kb of the region where the five EPAS1 genes are located in Han Chinese GWAS.

Based on the results of gene-based analysis, this work conducted a PPI study on Tibetan and Han Chinese patients. The results revealed a network centered on *LPIN1* in Cluster1 that connected *PLA2G4C*, *CEPT1*, *PPAPDC1B* and other genes (**Fig. 3A(b)**). These genes were enriched in phospholipid biosynthesis and metabolism as well as ammonium ion metabolism (P<0.05, Counts_gene_> 5, **Fig. 3A(b)**). Moreover, in Cluster2 a network with IL1B as an important center was identified. This network linked *EPAS1*, *HIF1A*, *HIF13A*, *ALOX15*, *MMP25*, and other genes (**Fig. 3A(b)**), and they were enriched in response to oxidative stress, regulation of apoptotic signaling pathways, steroid metabolic processes and alcohol metabolic processes (P<0.05, Counts_gene_>5, **Fig. 3A(b)**). In addition, this work identified four clusters in the Han PPI network (**Fig. 3A(c)**), and the pathway enrichment study (P<0.05, Counts_gene_>2, **Fig. 3A(b)**) suggested that Cluster1 was mainly related to the regulation of cell morphogenesis and differentiation, Cluster2 was mainly focused on the regulation of the nervous system, Cluster3 was mainly related to ion transport, and Cluster4 was focused on extracellular matrix organization and actin cytoskeleton reorganization-related pathways.

HAPC is a disease that does not adapt to low-pressure and hypoxic environments. Gene sets were compared between the genes with the differential VARIANTs identified in the GWAS (P<0.01) and the “hallmark Hypoxia” gene **(Table S6)** in the MSigDB database, which identified 38 and 16 intersecting genes in Tibetan and Han patients (**Fig. 3B(c)**). Based on 38 and 16 genes set GO analyze, common biological processes in response to hypoxia were identified between Tibetan and Han patients, including the “monosaccharide metabolic process” and “hexose metabolic process.” In addition, this study focused on the significant differences between Tibetans and Han Chinese patients. In Tibetan samples, more genes were enriched in the oxygen-sensing pathway, mainly including *EPAS1*, *HIF1A*, and *HIF3A*, with *HIF3A* and *EPAS1* having highly significant levels in the GWAS fraction (P = 2.88E-08, P = 2.92E-06), but not in the Han Chinese population, which may be related to natural selection^8^. Moreover, unlike Tibetans, in Han Chinese patients we focused on the pathway of the organophosphate catabolic process. In general, the genetic susceptibility difference in the oxygen-sensing pathway may be an important reason for the difference in HAPC between Tibetans and Han Chinese.

### Exploration of *EPAS1* and HAPC

Our study also focused on the genetic susceptibility of *EPAS1* and Tibetan HAPC. LD analysis of five differential VARIANTs in *EPAS1* (rs1374749, rs10178633, rs11678465, rs1868092, and rs11689694) by Tibetan groups (n_HAPC_=445, n_Non-HAPC_=524) enabled the identification of several haplotype blocks associated with HAPC risk. First, the paired *t*-test suggested significant differences between the LD (D’) values of five VARIANTs in HAPC and non-HAPC patients (P = 0.02). Furthermore, the differences in all haplotypes in 5 VARIANTs between HAPC and non-HAPC patients were checked using chi-squared tests, and the results showed that “GGTAC” was the HAPC risk haplotype block (P=5.48E-09, OR=2.15, **Fig. 3B(b)**). The blood phenotypic difference analysis showed that the Tibetan HAPC population in the “AACGT” haplotype block was reflected by a low hemoglobin level (*P*<0.001, **Fig. 3C**), a high HCT level, and a low lymphocyte level (*P*<0.01, **Fig. 3C**). Therefore, the GGTAC haplotype block of *EPAS1* may influence the hemoglobin concentration, is involved in the occurrence of HAPC and is a risk haplotype for HAPC.

To explore the LD state of other sites in the five-VARIANT haplotype block in *EPAS1* gene, this study observed the 20-kb region (chr2, 46596433:46617182) where five VARIANTs were located in the GWAS sample, including 13 VARIANTs in Tibetan patients and nine VARIANTs in Han Chinese patients. In the Tibetan population, compared with the non-HAPC group, HAPC patients showed significant differences in the LD status of rs1374749, rs10178633 and rs794912 compared with other VARIANTs (P=0.02, **Fig. 3C**). In the 20-kb *EPAS1* study in Han Chinese, 3 VARIANTs (rs1374749, rs10178633, and rs7594912) and high LD with other VARIANTs were identified as possible risk factors for HAPC in Han Chinese (P=0.003, **Fig. 3D**), where VARIANT genotypes were significantly associated with multiple blood phenotypes (P<0.05, **Table S3. Fig. S1B**). In summary, the linkage status of the rs1374749 locus with other *EPAS1* loci may be an important HAPC prevalence factor.

### Molecular diagnostic markers of HAPC in Tibetan and Han Chinese

Based on genetic susceptibility studies and phenotypic exploration, we found 11 significantly different phenotypes between HAPC and non-HAPC, such as PLT, RDW.SD and P-LCR, besides the significant differences in HGB, HTC and RBC. Blood phenotypes are intuitive features for the diagnosis of HAPC, and we explored the diagnostic power of 11 phenotypes for HAPC by means of logistic regression. We found that the 3 phenotypes had an independent diagnostic power of 0.7 or higher for HAPC in both Tibetans and Han Chinese (**Fig 5A(a); B (a)**), of which PLT was the highest. After that, molecular models of HAPC were constructed for Tibetans and Han Chinese (Tibetan-HRPM and Han-HRPM) combining 3 phenotypes and genetic susceptibility factors, respectively. First, the 791 volunteers were included in the Tibetan study, including 338 HAPC patients, and the 5-fold cross validation found that the AUC of Tibetan-HRPM model could reach 81.5% (AUC_Mean Train_=0.815, **Fig 5A(c)**), in which PLT, rs1868092, rs2567705 had a high diagnostic contribution (**Fig 5A(b)**). The groups split by median values of feature values using the Tibetan-HRPM model had significant HGB differences (**Fig 5A(d)**). Second, 751 volunteers were included in the Han study, of which 330 were HAPC patients, and a 5-fold cross validation found up to 82.8% in Han-HRPM (AUC_Mean Train_=0.828, **Fig 5B(c)**), in which PLT, rs1030005, and rs7618658 had a high diagnostic contribution (**Fig 5B(b)**). Similarly, populations segmented by median values of eigenvalues using the Han-HRPM model had significant HGB differences (**Fig 5B(d)**). In addition, the study used a Nomogram approach to plot the HAPC clinical risk diagnostic scales for Tibetan-HRPM and Han-HRPM.

**Fig. 5.**
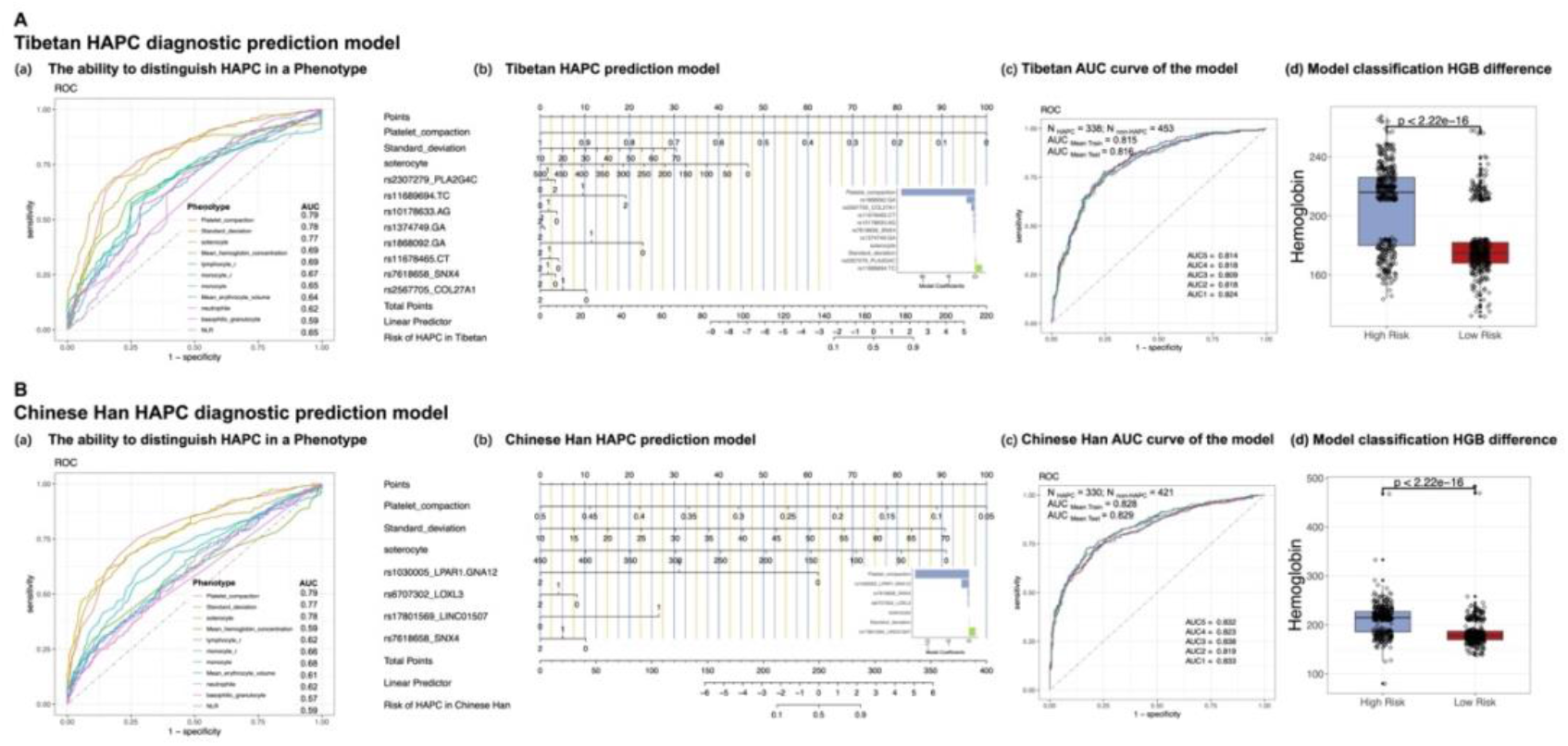
Molecular diagnostic markers of HAPC in Tibetan and Han Chinese: **A. (a).** The ROC curve of the diagnostic model of the HAPC between the Tibetan HAPC and the Non-HAPC. **(b).** Nomogram analysis of a combined molecular diagnostic model of Tibetan single phenotype and VARIANTs. **(c).** The 5-fold cross-validation ROC curve of the Tibetan combined molecular diagnostic model. **B. (a).** The ROC curve of the diagnostic model of the HAPC between the Chinese Han HAPC and the Non-HAPC. **(b).** Nomogram analysis of a combined molecular diagnostic model of Chinese Han single phenotype and VARIANTs. **(c).** The 5-fold cross-validation ROC curve of the Chinese Han combined molecular diagnostic model.

## Discussion

In this study, we identified 77 and 25 differential VARIANTs (P value < 5e-8) between Tibetan and Han Chinese HAPC and non-HAPC groups identified via GWAS analysis, respectively. Moreover, this study validated the common protective factor rs7618658 (C>T, *SNX4*) for Tibetan and Han HAPC patients. Furthermore, two missense variant sites were also suggested in Tibetans, among which rs2307279 (PLA2G4C, A38P) increased the stability of the pla24c protein (PolyPhen-2, I-Mutant2.0) in the Tibetan HAPC population. Afterward, at the macroscopic phenotype level, we found 13 blood characteristics were significantly different between HAPC and non-HAPC patients among Tibetans and Han Chinese. Remarkably, WBC and NEUT% differed between HAPC and non-HAPC patients in Tibetans and not in Han Chinese. Furthermore, this study found a significant correlation between HAPC susceptibility VARIANT and phenotypes differed between Tibetans and Han Chinese, for instance, rs7618658(SNX4) was significantly associated with MPV among Han Chinese, but not among Tibetans. The studies also explored the evolutionary genes of the oxygen-sensing pathway, and it was found that the GGTAC haplotype block of 5 strongly linked *EPAS1* loci in Tibetan patients had significantly increased HGB (P<0.05). Overall, our study suggested different genetic susceptibility in Tibetan and Han Chinese HAPC. The GO and PPI analyses suggested a possible difference in HAPC susceptibility between Tibetans and Han Chinese. Among them, the evolution of oxygen sensitivity of the oxygen-sensing pathway (*EPAS1*) in the Tibetan population was the main mechanism of HAPC disease. The susceptibility of Han Chinese to HAPC may stem from the ability to habituate to a hypoxic environment, such as increased angiogenesis and cell differentiation.

The blood phenotypic characteristics of Tibetan and Han Chinese HAPC patients have similarities and characteristics. Consistent with previous studies, Tibetan and Han Chinese HAPC patients had significant increases in RBC, HGB and HCT^11^. On this basis, this study also found differences between the blood phenotypes of Tibetan and Han Chinese HAPC patients, such as RBC (P<0.01), which were related to the adaptive evolution of the oxygen-sensing pathway (*EPAS1*)^37^. This work focused on the ability of these differential phenotypes to classify the HAPC population, where PLT, RDW.SD and P-LCR could reach an accuracy of more than 75% for the independent diagnosis of HAPC, suggesting a new diagnostic approach for HAPC. Studies have shown that RDW.SD may be regulated by the oxygen-sensing pathway (*HIF-1A*/*EPAS1*/*EPO*) in hypoxic environments^38–40^, and the reduction in PLT may also be related to the activation of the oxygen-sensing pathway (*HIF-1A*/*EPAS1*) downstream of the inflammatory response^16^. In addition, in a previous study a significant increase in erythrocytes in mice exposed to 500 ppm CO was accompanied by a significant decrease in PLT^41^, which is associated with hypoxic compensation. Therefore, PLT, RDW.SD, and P-LCR may be available as diagnostic standards for HAPC. This study also focused on clustering LYM% with RDW-SD specific to the Tibetan HAPC population. The diminished LYM% may represent a diminished immune system ^42, 43^, which is significantly associated with the five loci of *EPAS1* and may be related to the oxygen-sensing pathway in Tibetans. This investigation also found lower levels of NLR in Tibetans than in Han Chinese (P<0.001). NLR is associated with inflammation^44^, cardiovascular disease^45^, and thrombotic risk in true erythrocytosis^26^ and its elevation may cause atherosclerosis^45^. In general, the findings of this study suggest that Tibetan groups exposed to the evolution of oxygen-sensing pathways may have a lower state of immune activation as well as lower erythrocyte levels but may have an increased risk of complications due to HAPC, this possibility may need to be supported by more data.

This work focused on a missense risk variant site, rs2307279 (C>G, 5’UTR variant/missense variant(A38P), *PLA2G4C*, cPLA2-gamma, OR_combain_=1.02, Freq=0.1529), in the Tibetan patients. This site induces the substitution of the cPLA2-gamma protein amino acid 38th position A into proline and increases the stability of the protein tertiary structure (I-Mutant2.0). The database suggests that the GG genotype of rs2307279 significantly increases the mRNA expression of cPLA2 (P_GTEx_=1.3e-15). The cPLA2-gamma was activated in a hypoxic environment^46^ to affect the nuclear localization ability of *HIF-1A*^47^, which is involved in the hydrolysis of glycerophospholipids to produce free fatty acids and lysophospholipids^48^. In addition, cPLA2-gamma may cause the disruption of erythrocyte (RBC) membrane structure via damaging membrane integrity^49^, thus causing diminished RBC^50^ function or even hemolysis^51^, and ultimately erythropoiesis^52^. Other studies have shown that cPLA2-gamma inhibitors reduce the mRNA expression of *EPO*^47^. Based on the effect of cPLA2-gamma on HIF-1A as well as *EPO*, Tibetan groups carrying the rs2307279 variant GG genotype may be more susceptible to HAPC. In addition, this study also focused on the rs7618658 (C>T, *SNX4*) variant, which is a protective factor for HAPC. Its TT genotype was associated with higher NLR, NEUT, and MONO. Studies have shown that *SNX4* binds to phosphatidylinositol 3-phosphate (*PI3P*), contributes to direct cellular autophagy^53^ and is involved in the cyclic degradation of the transferrin receptor (TfnR)^54, 55^. Research has shown that *SNX4* is directly involved in cellular autophagy by binding to phosphatidylinositol 3-phosphate (*PI3P*)^53^. *SNX4* is involved in the cyclic degradation of the TfnR^54, 55^. Cellular autophagy may increase the ROS status, which activates the oxygen-sensing pathway^56, 57^. In summary, the combined genetic and phenotypic exploration of the Tibetan group revealed that the influence of Tibetan genetic susceptibility on oxygen-sensing pathways may have formed the unique blood phenotypic characteristics of Tibetan HAPC. The PPI network also showed the significant enrichment of genetic susceptibility to Tibetan HAPC in the phospholipid metabolism and oxygen-sensing pathways. Therefore, these findings suggest that the Tibetan HAPC population may have an evolutionary state that is more sensitive to oxygen-sensing pathways, such as the roles of the *EPAS1* and *PLA2G4C* genes in *HIF-1A*. However, the specific molecular mechanisms need to be explored experimentally at a later stage.

Different from Tibetans, no significant genetic susceptibility features affecting the oxygen perception pathway were identified in Han Chinese patients. The results indicate that Han Chinese HAPC genetic susceptibility may be associated with cell differentiation and angiogenesis. In the genetic exploration, the TT variant of rs1030005 (C>T, *LPAR1*/*GNA12*) significantly increased *GNG10* gene expression in visceral adipose tissue (GTEx, **Fig. S1B**, P=0.000040). The expression of *GNA12* by G protein subunit alpha 12 (Gα12) is involved in proteasomal ubiquitin-dependent proteolytic metabolism. *GNA12*- dependent Rho signaling has been shown to regulate the transcription factor AP-1 (activator protein-1)^58–, 60^, which mediates VEGF-induced endothelial cell migration and proliferation^61^. In the Han HAPC blood phenotype association study, rs1030005 (C>T, *LPAR1*/*GNA12*) had a TT genotype with lower RBC, HGB, HTC, BOSO, NEUT and Eoso. In general, this study found no genes associated with oxygen-sensing pathways in the Han population, and instead detected more angiogenesis and cell differentiation directions, such as *GNA12*. In addition, unlike Tibetans, the PPI network in the Han population suggested four possible pathway processes, namely the regulation of cell differentiation, vascular morphogenesis, ion transport, and cytoskeleton.

The genetic susceptibility exploration showed that sensitivity to oxygen-sensing pathways may be an important mechanism of disease in Tibetan HAPC. This sensitivity was mainly reflected in the secondary effects of *EPAS1* and *PLA2G4C* on *HIF-1A*. Increasing number of studies have shown that variants in the non-coding region of *EPAS1* are associated with reduced hemoglobin concentrations in Tibetans^62^. In addition, the LD-linked status of the *EPAS1* gene region is an important direction for plateau adaptation exploration^63^. Studies have shown that rs1447563 and rs4953388 variants in *EPAS1* are associated with hemoglobin concentration in healthy Tibetans^64^, which is inherited in linkage with rs1374749, rs1868092, and rs11678465^63^. Consistently, the present study showed that the AA variant in rs1374749 reflected lower hemoglobin in the Tibetan HAPC population. It was found that the 20-kb rs1374749 in the Tibetan and Han HAPC populations shared a common low-linkage feature with the status of other loci, and its importance as a variant that reduces hemoglobin concentration may be an important disease mechanism causing HAPC in Tibetans and Han Chinese. In addition, some studies have shown that the “GGG” haplotype block in the Han Chinese *EPAS1* population is a risk factor for HAPC^36^, but Tibetan-related haplotype studies have not reported a direct association with HAPC. The present study found that the GGTAC haplotype at five *EPAS1* loci was a risk haplotype for Tibetan HAPC, which was reflected in higher HGB levels. In general, the linkage genetics of *EPAS1* are an evolutionary feature of highland adaptation^63^; Han Chinese have not been subjected to this form of natural selection, and therefore the linkage status of *EPAS1* is at a lower level and is more likely to produce HAPC, which is consistent with the significantly higher HGB in Han Chinese populations compared to Tibetans. In contrast, the low chain of five *EPAS1* loci in the Tibetan HAPC population, especially the rs1374749 locus, may be a phenomenon of incomplete adaptation to the plateau, which implies an increase in the sensitivity of the population to oxygen perception as reflected by an increase in hemoglobin.

Although our study explored the genetic susceptibility factors of HAPC based on a relatively large sample size and obtained relatively new results, this study has limitations. The direction of the genetic susceptibility of the loci generated in the GWAS stage and subsequent validation stages may be biased.GWAS results showed that the rs2307279 (PLA2G4C) locus had a higher variant rate in HAPC, but after expanding the population, the variant rate was higher in the Non-HAPC population. Therefore, the specific genetic variants related to the genetic susceptibility of HAPC need to be verified in subsequent experiments.

## Author Contributions

**Zhuoma Basang:** Ideas; Conceptualization; Funding acquisition; Research plan; Writing. **Shixuan Zhang:** Ideas; Methodology; Writing. **Juan Wu:** Sample Collection; Programming ; Research plan. **Jiucun Wang:** Methodology; Programming; Research plan. **Yi Li:** Methodology; Programming; **Juan Wu:** Sample Collection; Research plan. **Xianwei Ke:** Sample Collection; Research plan. **Zhuoma Duoji:** Sample Collection; Research plan. **La Yang:** Sample Collection; Research plan. **Danzeng Qiangba:** Sample Collection. **Yang De:** Sample Collection. **Zixin Hu:** Writing-original draft. **Meng Hao:** Writing-original draft. **Yanyun Ma:** Writing-original draft.

## Data Availability

The data used in this study are available from the corresponding author upon reasonable request. The datasets generated and/or analyzed during the current study are not publicly available due to reason for data restrictions.

## Acknowledgments

We are most grateful to all the individuals who participated in this study, especially the Physical Examination Center, Fukang Hospital for their contributions to this study. This work was supported by grants from: The Science and Technology Department of Tibet (08080002), 2019 School-level Cultivation Project of Tibet University (ZDTSJH19-08), and Special Funds from the Central Finance to Support the Development of Local Universities [2018] No. 54; [2019] No. 1-19; [2020] No.79; [2021] No.1; (00060695/003). This research was supported by Shanghai Municipal Science and Technology Major Project (2017SHZDZX01), National Science Foundation of China (32288101, 32200536), and CAMS Innovation Fund for Medical Sciences (2019-I2M-5-066). The funders had no role in study design, data collection and analysis, decision to publish, or preparation of the manuscript. Finally, we thank LetPub (www.letpub.com) for its linguistic assistance during the preparation of this manuscript.

## Reference

1 León-Velarde, F. et al. Consensus statement on chronic and subacute high altitude diseases. High Alt Med Biol 6, 147–157, doi:10.1089/ham.2005.6.147 (2005).

2 Guan, W. et al. Sleep disturbances in long-term immigrants with chronic mountain sickness: a comparison with healthy immigrants at high altitude. Respir Physiol Neurobiol 206, 4–10, doi:10.1016/j.resp.2014.11.007 (2015).

3 Julian, C. G. et al. Perinatal hypoxia increases susceptibility to high-altitude polycythemia and attendant pulmonary vascular dysfunction. Am J Physiol Heart Circ Physiol 309, H565–573, doi:10.1152/ajpheart.00296.2015 (2015).

4 Jefferson, J. A. et al. Hyperuricemia, hypertension, and proteinuria associated with high-altitude polycythemia. Am J Kidney Dis 39, 1135–1142, doi:10.1053/ajkd.2002.33380 (2002).

5 Chen, S. [Investigation on the incidence of high altitude polycythemia and its hemoglobin characteristics in a Tibetan population]. Zhongguo Yi Xue Ke Xue Yuan Xue Bao 14, 237–243 (1992).

6 Painschab, M. S. et al. Association between serum concentrations of hypoxia inducible factor responsive proteins and excessive erythrocytosis in high altitude Peru. High Alt Med Biol 16, 26–33, doi:10.1089/ham.2014.1086 (2015).

7 Jiang, C. et al. Chronic mountain sickness in Chinese Han males who migrated to the Qinghai-Tibetan plateau: application and evaluation of diagnostic criteria for chronic mountain sickness. BMC Public Health 14, 701, doi:10.1186/1471-2458-14-701 (2014).

8 Lu, D. et al. Ancestral Origins and Genetic History of Tibetan Highlanders. Am J Hum Genet 99, 580–594, doi:10.1016/j.ajhg.2016.07.002 (2016).

9 Buroker, N. E. et al. EPAS1 and EGLN1 associations with high altitude sickness in Han and Tibetan Chinese at the Qinghai-Tibetan Plateau. Blood Cells Mol Dis 49, 67–73, doi:10.1016/j.bcmd.2012.04.004 (2012).

10 Xu, S. et al. A genome-wide search for signals of high-altitude adaptation in Tibetans. Mol Biol Evol 28, 1003–1011, doi:10.1093/molbev/msq277 (2011).

11 Fandrey, J. High-altitude polycythemia. Haematologica 90, 1 (2005).

12 Novy, M. J., Edwards, M. J. & Metcalfe, J. Hemoglobin Yakina. II. High blood oxygen affinity associated with compensatory erythrocytosis and normal hemodynamics. J Clin Invest 46, 1848–1854, doi:10.1172/jci105675 (1967).

13 Maran, J. & Prchal, J. Polycythemia and oxygen sensing. Pathol Biol (Paris) 52, 280–284, doi:10.1016/j.patbio.2004.02.006 (2004).

14 Lee, F. S. & Percy, M. J. The HIF pathway and erythrocytosis. Annu Rev Pathol 6, 165–192, doi:10.1146/annurev-pathol-011110-130321 (2011).

15 Liu, H. et al. EPAS1 regulates proliferation of erythroblasts in chronic mountain sickness. Blood Cells Mol Dis 84, 102446, doi:10.1016/j.bcmd.2020.102446 (2020).

16 McGettrick, A. F. & O’Neill, L. A. J. The Role of HIF in Immunity and Inflammation. Cell Metab 32, 524–536, doi:10.1016/j.cmet.2020.08.002 (2020).

17 Wu, M. F. et al. Hif-2α regulates lipid metabolism in alcoholic fatty liver disease through mitophagy. Cell Biosci 12, 198, doi:10.1186/s13578-022-00889-1 (2022).

18 Castelli, S., Ciccarone, F., Tavian, D. & Ciriolo, M. R. ROS-dependent HIF1α activation under forced lipid catabolism entails glycolysis and mitophagy as mediators of higher proliferation rate in cervical cancer cells. J Exp Clin Cancer Res 40, 94, doi:10.1186/s13046-021-01887-w (2021).

19 Rattner, A., Williams, J. & Nathans, J. Roles of HIFs and VEGF in angiogenesis in the retina and brain. J Clin Invest 129, 3807–3820, doi:10.1172/jci126655 (2019).

20 Zhao, Y. et al. Associations of high altitude polycythemia with polymorphisms in EPAS1, ITGA6 and ERBB4 in Chinese Han and Tibetan populations. Oncotarget 8, 86736–86746, doi:10.18632/oncotarget.21420 (2017).

21 Fan, X. et al. Associations of high-altitude polycythemia with polymorphisms in PIK3CD and COL4A3 in Tibetan populations. Hum Genomics 12, 37, doi:10.1186/s40246-018-0169-z (2018).

22 Chen, Y., Jiang, C., Luo, Y., Liu, F. & Gao, Y. Interaction of CARD14, SENP1 and VEGFA polymorphisms on susceptibility to high altitude polycythemia in the Han Chinese population at the Qinghai-Tibetan Plateau. Blood Cells Mol Dis 57, 13–22, doi:10.1016/j.bcmd.2015.11.005 (2016).

23 Gesang, L., Gusang, L., Dawa, C., Gesang, G. & Li, K. Whole-Genome Sequencing Identifies the Egl Nine Homologue 3 (egln3/phd3) and Protein Phosphatase 1 Regulatory Inhibitor Subunit 2 (PPP1R2P1) Associated with High-Altitude Polycythemia in Tibetans at High Altitude. Dis Markers 2019, 5946461, doi:10.1155/2019/5946461 (2019).

24 Adzhubei, I., Jordan, D. M. & Sunyaev, S. R. Predicting functional effect of human missense variants using PolyPhen-2. Curr Protoc Hum Genet Chapter 7, Unit7.20, doi:10.1002/0471142905.hg0720s76 (2013).

25 Hao, M. et al. The HuaBiao project: whole-exome sequencing of 5000 Han Chinese individuals. J Genet Genomics 48, 1032–1035, doi:10.1016/j.jgg.2021.07.013 (2021).

26 Carobbio, A. et al. Neutrophil-to-lymphocyte ratio is a novel predictor of venous thrombosis in polycythemia vera. Blood Cancer J 12, 28, doi:10.1038/s41408-022-00625-5 (2022).

27 Canela-Xandri, O., Rawlik, K. & Tenesa, A. An atlas of genetic associations in UK Biobank. Nat Genet 50, 1593–1599, doi:10.1038/s41588-018-0248-z (2018).

28 Consortium, G. T. The Genotype-Tissue Expression (GTEx) project. Nat Genet 45, 580–585, doi:10.1038/ng.2653 (2013).

29 Jumper, J. et al. Highly accurate protein structure prediction with AlphaFold. Nature 596, 583–589, doi:10.1038/s41586-021-03819-2 (2021).

30 Capriotti, E., Fariselli, P. & Casadio, R. I-Mutant2.0: predicting stability changes upon variant from the protein sequence or structure. Nucleic Acids Res 33, W306–310, doi:10.1093/nar/gki375 (2005).

31 Pejaver, V. et al. Inferring the molecular and phenotypic impact of amino acid variants with MutPred2. Nat Commun 11, 5918, doi:10.1038/s41467-020-19669-x (2020).

32 Courvoisier, D. S., Combescure, C., Agoritsas, T., Gayet-Ageron, A. & Perneger, T. V. Performance of logistic regression modeling: beyond the number of events per variable, the role of data structure. J Clin Epidemiol 64, 993–1000, doi:10.1016/j.jclinepi.2010.11.012 (2011).

33 Li, Y. et al. knnAUC: an open-source R package for detecting nonlinear dependence between one continuous variable and one binary variable. BMC Bioinformatics 19, 448, doi:10.1186/s12859-018-2427-4 (2018).

34 Li, Y. et al. Using Composite Phenotypes to Reveal Hidden Physiological Heterogeneity in High-Altitude Acclimatization in a Chinese Han Longitudinal Cohort. Phenomics 1, 3–14, doi:10.1007/s43657-020-00005-8 (2021).

35 Chandrasekhar, C., Pasupuleti, S. K. & Sarma, P. Novel variants in the EPO-R, VHL and EPAS1 genes in the Congenital Erythrocytosis patients. Blood Cells Mol Dis 85, 102479, doi:10.1016/j.bcmd.2020.102479 (2020).

36 Chen, Y., Jiang, C., Luo, Y., Liu, F. & Gao, Y. An EPAS1 haplotype is associated with high altitude polycythemia in male Han Chinese at the Qinghai-Tibetan plateau. Wilderness Environ Med 25, 392–400, doi:10.1016/j.wem.2014.06.003 (2014).

37 Xu, J. et al. EPAS1 Gene Polymorphisms Are Associated With High Altitude Polycythemia in Tibetans at the Qinghai-Tibetan Plateau. Wilderness Environ Med 26, 288–294, doi:10.1016/j.wem.2015.01.002 (2015).

38 Yčas, J. W. Toward a Blood-Borne Biomarker of Chronic Hypoxemia: Red Cell Distribution Width and Respiratory Disease. Adv Clin Chem 82, 105–197, doi:10.1016/bs.acc.2017.06.002 (2017).

39 Yčas, J. W., Horrow, J. C. & Horne, B. D. Persistent increase in red cell size distribution width after acute diseases: A biomarker of hypoxemia? Clin Chim Acta 448, 107–117, doi:10.1016/j.cca.2015.05.021 (2015).

40 Emans, M. E. et al. Determinants of red cell distribution width (RDW) in cardiorenal patients: RDW is not related to erythropoietin resistance. J Card Fail 17, 626–633, doi:10.1016/j.cardfail.2011.04.009 (2011).

41 Penney, D. G. & Bishop, P. A. Hematologic changes in the rat during and after exposure to carbon monoxide. J Environ Pathol Toxicol 2, 407–415 (1978).

42 Huang, H. et al. Lymphocyte percentage as a valuable predictor of prognosis in lung cancer. J Cell Mol Med 26, 1918–1931, doi:10.1111/jcmm.17214 (2022).

43 Schmidt, H. et al. Elevated neutrophil and monocyte counts in peripheral blood are associated with poor survival in patients with metastatic melanoma: a prognostic model. Br J Cancer 93, 273–278, doi:10.1038/sj.bjc.6602702 (2005).

44 Ekinci, O. & Ekinci, A. The connections among suicidal behavior, lipid profile and low-grade inflammation in patients with major depressive disorder: a specific relationship with the neutrophil-to-lymphocyte ratio. Nord J Psychiatry 71, 574–580, doi:10.1080/08039488.2017.1363285 (2017).

45 Wang, H. et al. The relationship between neutrophil to lymphocyte ratio and artery stiffness in subtypes of hypertension. J Clin Hypertens (Greenwich) 19, 780–785, doi:10.1111/jch.13002 (2017).

46 Lambert, I. H., Pedersen, S. F. & Poulsen, K. A. Activation of PLA2 isoforms by cell swelling and ischaemia/hypoxia. Acta Physiol (Oxf) 187, 75–85, doi:10.1111/j.1748-1716.2006.01557.x (2006).

47 Osada-Oka, M., Takahashi, M., Akiba, S. & Sato, T. Involvement of Ca2+-independent phospholipase A2 in the translocation of hypoxia-inducible factor-1alpha to the nucleus under hypoxic conditions. Eur J Pharmacol 549, 58–62, doi:10.1016/j.ejphar.2006.08.026 (2006).

48 Yamashita, A. et al. Subcellular localization and lysophospholipase/transacylation activities of human group IVC phospholipase A2 (cPLA2gamma). Biochim Biophys Acta 1791, 1011–1022, doi:10.1016/j.bbalip.2009.05.008 (2009).

49 Wu, X. Y. et al. [Study on the relationship between cytosolic phospholipase A2-gamma activation and myocardial cell injury during cardiopulmonary bypass]. Zhongguo Wei Zhong Bing Ji Jiu Yi Xue 17, 417–420 (2005).

50 Kostara, C. E., Tsiafoulis, C. G., Bairaktari, E. T. & Tsimihodimos, V. Altered RBC membrane lipidome: A possible etiopathogenic link for the microvascular impairment in Type 2 diabetes. J Diabetes Complications 35, 107998, doi:10.1016/j.jdiacomp.2021.107998 (2021).

51 Díaz, C. et al. Modulation of the susceptibility of human erythrocytes to snake venom myotoxic phospholipases A(2): role of negatively charged phospholipids as potential membrane binding sites. Arch Biochem Biophys 391, 56–64, doi:10.1006/abbi.2001.2386 (2001).

52 Yao, H., Ma, Y. & Huang, L. J. Deletion of miR-451 curbs JAK2(V617F)-induced erythrocytosis in polycythemia vera by oxidative stress-mediated erythroblast apoptosis and hemolysis. Haematologica 105, e153–e156, doi:10.3324/haematol.2018.210799 (2020).

53 Ravussin, A., Brech, A., Tooze, S. A. & Stenmark, H. The phosphatidylinositol 3-phosphate-binding protein SNX4 controls ATG9A recycling and autophagy. J Cell Sci 134, doi:10.1242/jcs.250670 (2021).

54 Sakane, H. et al. α-Taxilin interacts with sorting nexin 4 and participates in the recycling pathway of transferrin receptor. PLoS One 9, e93509, doi:10.1371/journal.pone.0093509 (2014).

55 Traer, C. J. et al. SNX4 coordinates endosomal sorting of TfnR with dynein-mediated transport into the endocytic recycling compartment. Nat Cell Biol 9, 1370–1380, doi:10.1038/ncb1656 (2007).

56 Dizin, E. et al. Activation of the Hypoxia-Inducible Factor Pathway Inhibits Epithelial Sodium Channel-Mediated Sodium Transport in Collecting Duct Principal Cells. J Am Soc Nephrol 32, 3130–3145, doi:10.1681/asn.2021010046 (2021).

57 Zhang, T. et al. Alpinetin inhibits breast cancer growth by ROS/NF-κB/HIF-1α axis. J Cell Mol Med 24, 8430–8440, doi:10.1111/jcmm.15371 (2020).

58 Krakstad, B. F., Ardawatia, V. V. & Aragay, A. M. A role for Galpha12/Galpha13 in p120ctn regulation. Proc Natl Acad Sci U S A 101, 10314–10319, doi:10.1073/pnas.0401366101 (2004).

59 Suzuki, N., Nakamura, S., Mano, H. & Kozasa, T. Galpha 12 activates Rho GTPase through tyrosine-phosphorylated leukemia-associated RhoGEF. Proc Natl Acad Sci U S A 100, 733–738, doi:10.1073/pnas.0234057100 (2003).

60 Tateiwa, K., Katoh, H. & Negishi, M. Socius, a novel binding partner of Galpha12/13, promotes the Galpha12-induced RhoA activation. Biochem Biophys Res Commun 337, 615–620, doi:10.1016/j.bbrc.2005.09.097 (2005).

61 Jia, J. et al. AP-1 transcription factor mediates VEGF-induced endothelial cell migration and proliferation. Microvasc Res 105, 103–108, doi:10.1016/j.mvr.2016.02.004 (2016).

62 Yi, X. et al. Sequencing of 50 human exomes reveals adaptation to high altitude. Science 329, 75–78, doi:10.1126/science.1190371 (2010).

63 Lou, H. et al. A 3.4-kb Copy-Number Deletion near EPAS1 Is Significantly Enriched in High-Altitude Tibetans but Absent from the Denisovan Sequence. Am J Hum Genet 97, 54–66, doi:10.1016/j.ajhg.2015.05.005 (2015).

64 Hanaoka, M. et al. Genetic variants in EPAS1 contribute to adaptation to high-altitude hypoxia in Sherpas. PLoS One 7, e50566, doi:10.1371/journal.pone.0050566 (2012).

